# Progression and prediction of sensorineural hearing loss in older adults using a convolutional neural network model and Markov chains

**DOI:** 10.64898/2026.11.23.26368371

**Authors:** Hector Gabriel Corrale de Matos, Sarah Rodrigues Thanis Garrido, Kátia de Freitas Alvarenga, Kelton Augusto Pontara da Costa, Lilian Cássia Bórnia Jacob

## Abstract

**Objectives:** This study compared a convolutional neural network (CNN) and linear regression for predicting mean hearing thresholds at the last audiogram and their variation from the first audiogram. It used audiometric and clinical data from older adults with bilateral sensorineural hearing loss. Markov chains estimated hearing-loss worsening and stability probabilities between assessments. Analyses considered hypertension, diabetes, occupational noise exposure and age.

**Design:** This retrospective study analysed serial audiograms and clinical records. Data included 823 adults with bilateral sensorineural hearing loss. Predictive analyses included 394 complete-case records, with 79 records per ear in the independent test set. Cross-validation selected model configurations and evaluated predictive performance across data partitions. Markov chains estimated worsening and stability probabilities across hearing-loss severity categories.

**Results:** CNN achieved cross-validation mean absolute errors of 6.475 dB HL (right ear, RE) and 6.047 dB HL (left ear, LE) and linear regression registered 6.054 dB HL (RE) and 6.170 dB HL (LE). CNN (independent evaluation) showed significantly lower prediction error in the LE after Holm correction (*p*_adj_ = 0.0328). No significant advantage was observed in the RE (*p*_adj_ = 0.1969). Markov state-averaged conditional worsening probabilities were 34.0% (RE) and 35.3% (LE). Observed worsening proportions were 42.2% (RE; 347/823) and 40.9% (LE; 337/823). Corresponding stability probabilities were 57.8% (RE) and 58.6% (LE). No statistically significant between-group differences were observed for occupational noise exposure, hypertension, or diabetes. Worsening estimates increased across age groups.

**Conclusions:** This study provides conditional probabilities of hearing-loss worsening and stability in older adults. CNN predictions and Markov estimates suggest that threshold prediction and severity transitions offer complementary descriptions of hearing changes. The ear-specific CNN advantage warrants further investigation. This characterisation may support assessment of changes in hearing thresholds alongside progression to more severe hearing-loss categories.

## 1 Introduction

Age-related hearing loss affects a significant proportion of the population (Li et al., 2022). Hearing loss affects communication and social participation and it does not remain stable throughout life (World Health Organization, 2021; Cruickshanks et al., 2020). Cross-sectional modelling has suggested that hearing deterioration accelerates with age (Sharma et al., 2021). Longitudinal observations have also documented changes in hearing thresholds in older adults (Göthberg et al., 2019). Serial audiometric assessments allow clinicians to monitor hearing-threshold levels and changes between successive evaluations (Dillard, Ghosh, et al., 2025; Vaden et al., 2017). Clinical and environmental factors are also associated with hearing loss and its progression over time (Dillard, Humes, et al., 2025; Linssen et al., 2014). Prior data found an association between high blood pressure and greater hearing loss over a period of approximately three years (Mick et al., 2023). In addition, published research associated diabetes with poorer hearing in cross-sectional analyses (Mick et al., 2023). Another study reported an association between occupational noise exposure and a higher prevalence of moderate to severe hearing loss in adults (Gopinath et al., 2021). Indeed, it also identified a higher incidence of hearing loss over a ten-year observation period (Gopinath et al., 2021). Nevertheless, reported associations between clinical and environmental factors and progression of age-related hearing loss vary across populations and study designs. Longitudinal approaches may support examination of the role of clinical and environmental factors in individual hearing trajectories over time.

In parallel, machine learning is being incorporated into audiology to extract complex patterns from audiometric and clinical data (Wasmann et al., 2022). Machine learning approaches have been applied to estimate audiometric thresholds and hearing status using audiometric, sample characteristics, and clinical features (Barbour et al., 2019; Liu et al., 2021; Liu & Gong, 2024; Song et al., 2015; Zhao et al., 2019). In these examples, models were designed to estimate or characterise the hearing level observed at assessment. Other machine learning approaches have predicted hearing thresholds from demographic, clinical and subjective information (Gathman et al., 2023). Machine learning has also been used to classify types of hearing loss from pure-tone audiometry data (Kassjański et al., 2024). These applications primarily aimed to characterise current hearing status (Gathman et al., 2023; Kassjański et al., 2024). Beyond these applications, machine learning has also been applied to longitudinal audiometric data to predict future changes in hearing thresholds. For example, a study used previous audiometric measurements to predict subsequent hearing thresholds in individuals with gene-related sensorineural hearing loss (Chen et al., 2024).

Nevertheless, longitudinal prediction in older adults with established bilateral sen-sorineural hearing loss remains under-explored. A second research question concerns estimating transition probabilities between hearing-loss severity categories. Such transitions may indicate worsening or stability between assessments. In this regard, longitudinal audiometric data have been used to characterise patterns of hearing change over time. These patterns included differences in stability and progression of audiometric profiles (Chen et al., 2024; Vaden et al., 2017). These longitudinal analyses, however, primarily treat variation over time as a change in audiometric thresholds or hearing status. Furthermore, these studies did not estimate probabilities of deterioration or stability between severity categories conditional on clinical factors.

Therefore, the primary objective was to compare a convolutional neural network and linear regression in predicting mean hearing threshold and its variation, based on audiometric and clinical data from older adults with bilateral sensorineural hearing loss. The secondary objective was to use Markov chains to estimate probabilities of hearing loss worsening or remaining stable between assessments, according to hypertension, diabetes, occupational noise exposure, and age.

## 2 Materials and Methods

### 2.1 Study Design and Setting

This retrospective longitudinal study included records from the Hearing Health Service at Bauru School of Dentistry, University of São Paulo. Clinical records covered 2008 to March 2023. First and last available audiograms defined each participant’s observation interval. Intervals were recorded in completed years rather than calculated from differences between mean ages at first and last audiograms. Research Ethics Committee approval was granted (Protocol No. 1,745,659). STROBE recommendations were followed (von Elm et al., 2007).

### 2.2 Study Population

The study analysed 823 records from patients with bilateral sensorineural hearing loss who underwent audiometric evaluation. The database comprised analyses of 1,646 audiograms. The study included only records of bilateral sensorineural hearing loss (four-frequency pure-tone average *≥* 20 dB HL) World Health Organization (2021). Mean age at the first audiogram was 70.5 years (SD 8.1; n = 823), and mean age at the last audiogram was 77.0 years (SD 8.1; n = 823). Age at the first audiogram, which ranged from 46 to 93 years, defined Markov age groups. The sample included 396 males (48.1%) and 427 females (51.9%). Patients with conductive or mixed hearing loss, chronic otitis media, otosclerosis, ototoxic medication use, previous otologic surgery, or acute acoustic trauma did not meet selection criteria.

### 2.3 Audiological Record Annotation and Data Collection

Data collection involved extracting audiological information from patient records and organising it in an Excel spreadsheet. For each patient, the database included right- and left-ear air-conduction thresholds at 250, 500, 1000, 2000, 3000, 4000, 6000, and 8000 Hz from the first and last audiological evaluations. It also included the four-frequency pure-tone average (PTA_4_) for each ear and evaluation, as well as the date of each evaluation. Clinical variables included age, sex, occupational noise exposure, tinnitus, hypertension, and diabetes. We also recorded other comorbidities reported in these records. Hypertension, diabetes, and occupational noise exposure were coded from information documented in clinical records. Age was summarised with mean, standard deviation, median, interquartile range, and minimum– maximum values. Sample characteristics were compared by sex using Pearson’s *χ*^2^ test for categorical variables and Welch’s *t*-test for continuous variables.

### 2.4 Data Cleaning and Preprocessing

The analysis pipeline standardised the dataset before computational modelling. Preprocessing converted fields to numeric format. It also addressed invalid and missing entries, standardised categorical labels, and capped audiometric thresholds at 120 dB HL. Subsequently, analysts classified the hearing loss level for each ear based on PTA_4_. This followed the World Health Organization (2021) classification. The preprocessing procedure did not impute missing clinical covariates; predictive analyses used records with complete clinical covariates. Tinnitus categories (unilateral right, unilateral left and bilateral) were grouped as tinnitus presence for prediction; missing values were retained.

### 2.5 Missing Audiometric Data

Data processing automatically excluded observations with missing values in variables required by computational models other than audiometric frequencies from the first audiogram. A masked-autoencoder imputer was fitted separately within each training partition using frequencies from the first audiogram. Validation and test observations were transformed without refitting the imputer. The autoencoder used fully connected layers with 64 and 32 neurons in the encoder and 64 neurons followed by the original input dimension in the decoder. It used rectified linear unit activations and mean squared error loss. Before training, preprocessing replaced missing entries with zero and standardised inputs. Adam optimisation (Kingma & Ba, 2015) used a learning rate of 1 *×* 10*^−^*^3^, and the model trained for 50 epochs before reconstructing missing values, which preprocessing then transformed back to the original measurement scale. When a training partition had no missing frequencies, missing values in held-out observations were filled with training-column medians. Only audiometric predictors from the first audiogram were imputed; outcomes and clinical covariates remained unimputed.

### 2.6 Predictive Outcome and Clinical Interpretation

The primary outcome was the mean hearing threshold at the last audiogram. We expressed this outcome in dB HL and modelled each ear separately. Predictors included air-conduction thresholds from the first audiogram at 250, 500, 1000, 2000, 3000, 4000, 6000, and 8000 Hz, age at the first audiogram, sex, hypertension, diabetes, occupational noise exposure, and binary tinnitus presence. Predictive analyses included tinnitus as a clinically relevant symptom accompanying sensorineural hearing loss. Its inclusion allowed assessment of its predictive contribution alongside audiometric and other clinical variables. CNN and linear regression analyses used 394 records with complete clinical covariates. Markov analyses retained all 823 records with hearing-loss severity grades. Continuous regression modelled each target, with higher predicted dB HL values indicating poorer future hearing. Two target formulations were evaluated. One predicted mean hearing threshold at the last audiogram directly, whereas the other predicted change from the first audiogram. We defined this change as the threshold at the last audiogram minus the threshold at the first audiogram. For the model predicting threshold change, the predicted variation was added to the first-audiogram threshold to estimate the threshold at the last audiogram.

### 2.7 Convolutional Neural Network

The analysis employed a compact one-dimensional convolutional neural network (1D-CNN). It used separate convolutional and fully connected branches for audiometric and clinical predictors. This CNN model is a type of neural network that uses convolutional layers (layers that apply learnable filters to local segments of the input) to learn patterns and relationships within input data. We chose this architecture because the convolutional branch can identify relevant local patterns and interactions among data.

The CNN was applied to audiometric thresholds, while the fully connected branch incorporated clinical predictors into the model. This combination allows the model to jointly use audiometric and clinical information for hearing threshold prediction. The CNN model is based on the following sequence of equations:

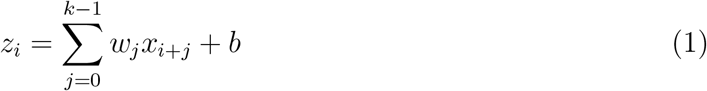

where *x* represents input audiometric thresholds, *w* represents the learnable convolutional filter, *k* is the filter size, and *b* is the bias term.

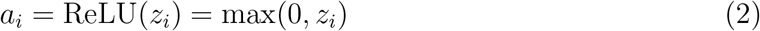

where ReLU represents the activation function that introduces nonlinearity into the model.

Resulting features are then combined with clinical predictors through the connected branch to generate the final prediction:

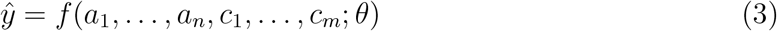

where *a*_1_*, …, a_n_* represent extracted audiometric features, *c*_1_*, …, c_m_* represent clinical predictors, *θ* represents model parameters, and *y*^ represents the predicted hearing threshold. For audiometric inputs, the convolutional branch received eight frequencies from the first audiogram (Figure 1). It divided data into two parallel pathways with kernel sizes of 3 and 5. Both pathways applied the same padding, batch normalisation, and rectified linear unit activation. Subsequently, a fusion convolution combined their outputs with a kernel size of 3. The fusion layer applied batch normalisation, rectified linear unit activation, and one-dimensional dropout. Adaptive average and maximum pooling summarised the fused representation. Next, the model concatenated pooled outputs and fed them into a 48-unit fully connected layer with rectified linear activation and dropout. In parallel, the clinical branch processed its predictors through a 16-unit fully connected layer with rectified linear activation and dropout. Thereafter, the model concatenated audiological and clinical representations. A 32-unit fully connected layer processed the combined representation with rectified linear activation and dropout. Then, a linear unit generated the continuous prediction.

**Figure 1:**
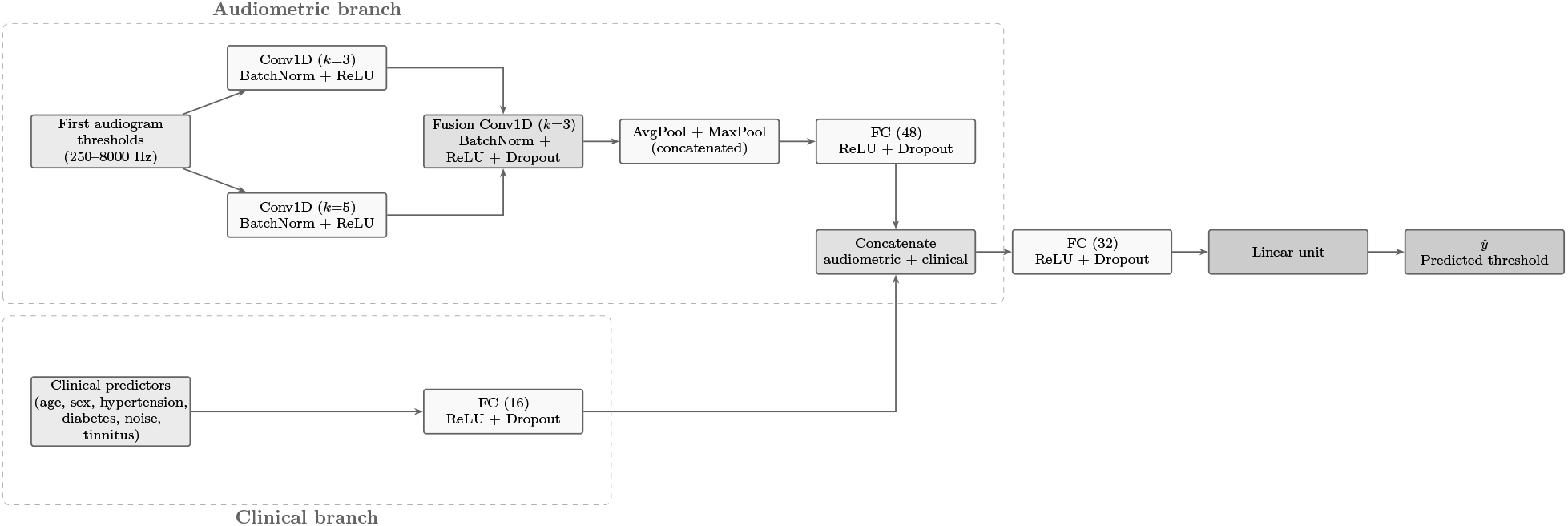
Architecture of the compact one-dimensional convolutional neural network (1D-CNN). The audiometric branch (top) applies two parallel convolutional pathways with kernel sizes 3 and 5, followed by a fusion convolution, adaptive average and maximum pooling, and a fully connected layer. The clinical branch (bottom) processes clinical predictors through a separate fully connected layer. Both representations are concatenated and passed through a final fully connected layer and a linear output unit to generate the predicted hearing threshold.

### 2.8 Hyperparameter Selection

We evaluated four predefined CNN configurations. These configurations varied channel sizes, dropout probability, learning rate, and weight decay. Channel configurations were (8, 16), (16, 16), and (16, 32), with dropout values of 0.20, 0.30, and 0.40, learning rates of 0.001 and 0.0005, and weight decay values of 10*^−^*^4^, 5 *×* 10*^−^*^4^, and 10*^−^*^3^. All configurations used kernel sizes of 3 and 5. Both direct-final and delta target formulations were evaluated. Mean absolute error defined the primary model-selection criterion.

### 2.9 Model Training

Model training involved standardising continuous audiometric predictors, clinical predictors, and target values with separate StandardScaler transformations (Pedregosa et al., 2011). Each scaler used only corresponding training data. Before evaluation, the model transformed predictions back to the original dB HL scale. Next, each training procedure allocated 15% of available training observations to internal validation. The validation split used a deterministic permutation based on the model seed. Mini-batches contained up to 32 observations, with a minimum batch size of four. AdamW optimisation (Loshchilov & Hutter, 2019) minimised mean squared error loss. During training, gradient clipping limited gradient norms to 1.0. Meanwhile, the learning-rate scheduler reduced the learning rate by a factor of 0.5 when validation loss failed to improve. It used a patience of six epochs and a minimum learning rate of 10*^−^*^5^. Training lasted for a maximum of 250 epochs, with early stopping after 20 epochs without sufficient improvement. An improvement greater than 10*^−^*^5^ in validation loss reset the stopping criterion. Afterwards, the procedure retained the model state with the lowest validation loss.

### 2.10 Nested Cross-Validation

The predictive analysis used nested cross-validation. In the outer loop, five folds comprised the validation structure, while the inner loop used three folds. Both loops shuffled observations and used fixed random seeds. Within each outer training set, the inner loop evaluated eight candidate combinations, representing four CNN configurations and two target formulations. It selected the candidate with the lowest MAE. Subsequently, the outer loop evaluated this candidate on the fold reserved for testing. Audiometric imputation and scaling were fitted only within training partitions and then applied to held-out observations. This nested procedure was repeated across 10 predefined seeds: 42, 123, 456, 789, 1011, 1337, 2023, 2024, 9999, and 11235.

### 2.11 Predictive Performance

We evaluated predictive performance using mean absolute error (MAE), root mean squared error (RMSE), and the coefficient of determination (*R*^2^). These metrics compare models in terms of prediction error and explained variance. MAE represents the average absolute difference between predicted and observed hearing thresholds:

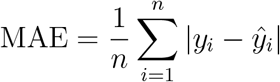

where *y_i_* represents the observed hearing threshold, *y*^*_i_* represents the predicted hearing threshold, and *n* represents the number of observations. RMSE gives greater weight to larger prediction errors:

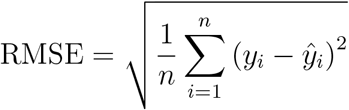

Both metrics were expressed in dB HL. *R*^2^ indicates the proportion of variability in observed hearing thresholds explained by the model:

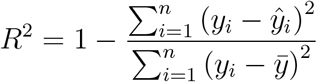

where *ȳ* represents the mean of observed hearing thresholds. It allows comparison of predictive performance across models. One performance estimate was calculated for each seed and ear. Mean performance and descriptive 2.5th–97.5th percentiles across seeds summarised variation across repeated partitions; these percentiles were not treated as population confidence intervals. As a reference comparison, linear regression used the same standardised inputs, outer folds, and seeds as the CNN. Seed-level CNN–regression differences were described without inferential testing. For held-out predictions, 2,000 participant-level bootstrap samples estimated the confidence interval for the mean paired absolute-error difference, and 9,999 paired sign-flip permutations provided a two-sided test. Cohen’s *d* was calculated as the mean paired difference in absolute error divided by the standard deviation of paired differences. Values of 0.2, 0.5, and 0.8 were used as reference thresholds for small, moderate, and large effects, respectively.

### 2.12 CNN Ensemble

A second CNN ensemble was generated on a fixed held-out test set. It allocated 20% of observations to testing and used random state 4242 for the train–test split. Test observations did not participate in hyperparameter selection. Meanwhile, the training subset underwent the same three-fold inner cross-validation procedure. This procedure selected one CNN configuration and one target formulation from the training subset. Subsequently, eight CNNs were trained with independent random seeds, and the ensemble averaged their predictions. For the delta formulation, each model predicted change from the first audiogram. The procedure then reconstructed the threshold at the last audiogram by adding the predicted change to the first-audiogram mean threshold. In contrast, the direct formulation predicted the last-audiogram threshold without reconstruction. For each test observation, the analysis calculated the standard deviation across eight predictions. A persistence comparator predicted the threshold at the last audiogram as equal to that at the first audiogram. It served as a clinical reference.

### 2.13 Markov Analysis

The secondary analysis evaluated transitions between hearing-loss severity states. A Markov chain is a probabilistic model that describes transitions between discrete states based on the current state. It was chosen to characterise changes in hearing-loss severity between evaluations and to quantify the probability of transitioning from one severity state to another. For each ear, hearing grades at the first and last audiograms defined transition states. Source severity codes 2 (mild), 3 (moderate), 4 (moderately severe), 5 (severe), 6 (profound) and 7 (complete) were mapped to states 0, 1, 2, 3, 4 and 4, respectively; the last state pooled profound and complete loss. Figure 2 illustrates the resulting state-transition structure. Transitions described the observed intervals between assessments (mean 6.0 years, SD 3.1), not annual rates.

**Figure 2:**
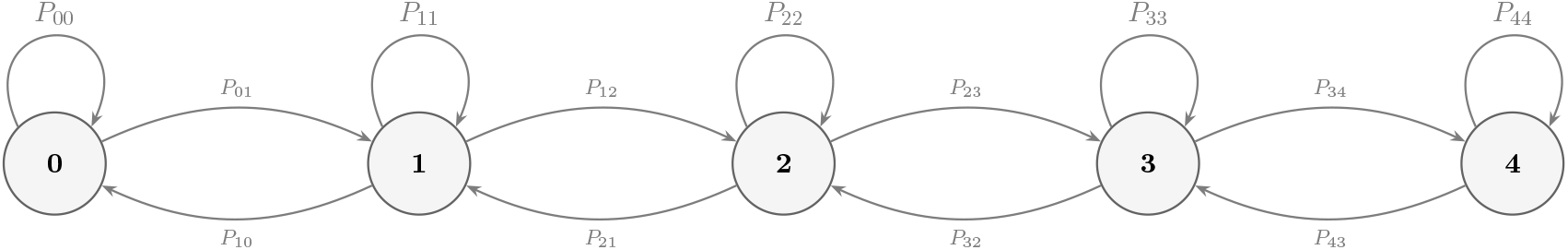
Schematic state-transition diagram for the five-state hearing-loss severity Markov chain. Nodes represent severity states (0 = mild to 4 = profound/complete). Source grades 6 and 7 are pooled in state 4. Curved arrows above states represent worsening transitions (upper-triangular entries, j > i); arrows below represent improvement transitions (j < i); self-loops represent the probability of remaining in the same severity state (diagonal entries, stability). For clarity, not all 20 off-diagonal transitions are shown; only adjacent-state transitions are illustrated as an example.

Next, the analysis constructed a 5 *×* 5 transition matrix for each ear. Rows represented starting severity states. Let *S_t_ ∈ {*0, 1, 2, 3, 4*}* denote the hearing-loss severity state at evaluation *t*, where 0 corresponds to mild and 4 corresponds to profound/complete hearing loss. The transition matrix *P* is defined as

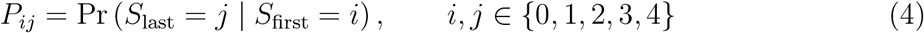

where *P_ij_*represents the probability of transitioning from starting state *i* (first audiogram) to state *j* (last audiogram). The analysis normalised each row by its row total and assigned zero values to rows without observations. The empirical transition matrix was estimated from observed counts as

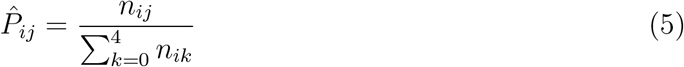

where *n_ij_* denotes the number of patients observed transitioning from state *i* to state *j*. Each row was normalised to satisfy the stochastic matrix constraint

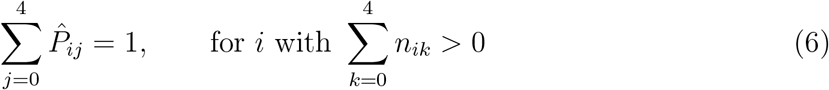

Subsequently, 2,000 participant-level bootstrap resamples stratified by starting severity state generated transition matrices and corresponding worsening and stability estimates. Worsening represented movement to a higher hearing-loss state. Then, worsening probability was defined as the unweighted mean of upper-triangular transition probabilities across five represented starting states:

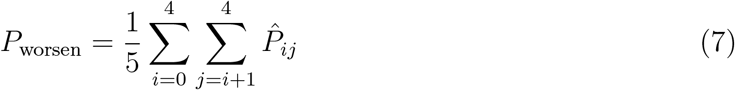

This measure represented the mean conditional probability of transition to a higher state and not the population-level marginal probability. The terminal state has no higher severity category, so its worsening probability is structurally zero. Observed worsening proportion was calculated separately as the number of participants with a higher severity state at the last audiogram divided by the total number assessed. Stability probability represented the mean of diagonal transition probabilities across five starting states:

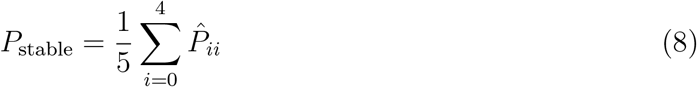

Bootstrap distributions provided percentile-based 95% confidence intervals. For each of *B* = 2,000 bootstrap resamples, a transition matrix *P*^^(^*^b^*) was recomputed, and corresponding 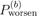 and 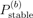 were calculated using Equations 7 and 8. Bootstrap 95% confidence intervals were obtained from the 2.5th and 97.5th percentiles of resulting distributions:

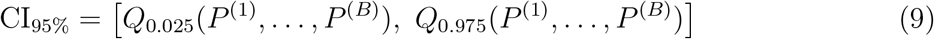

where *Q_α_*(*·*) denotes the empirical *α*-quantile of the bootstrap distribution.

For conditional Markov analysis, age at the first audiogram defined three groups: *<* 70 years, 70–79 years, and *≥* 80 years. Hypertension, diabetes, occupational noise exposure, and age group defined subgroup analyses. Each subgroup required at least 30 observations. Within each ear, the analysis compared presence and absence of hypertension, diabetes, and occupational noise exposure. The between-group estimand was the unweighted mean of within-state differences in worsening probability over starting states represented in both groups. Two thousand participant-level bootstrap resamples within initial states provided 95% confidence intervals for group differences. Two-sided permutation tests (4,999 permutations), shuffling group labels within starting states, provided corresponding *p*-values. No bootstrap-standardised effect size was calculated.

For formal hypothesis testing, Holm–Bonferroni correction was applied separately within two hypothesis families: two held-out CNN ensemble versus linear-regression comparisons (one per ear) and six Markov subgroup comparisons (hypertension, diabetes, and occupational noise exposure in each ear). An adjusted significance threshold of 0.05 was used.

### 2.14 Clinical Model Interpretation

Clinical interpretation focused on mean hearing thresholds at the last audiogram. Thus, the model estimated that threshold from first-audiogram data and clinical characteristics. Higher predicted dB HL values indicated poorer hearing. Feature interpretation used permutation feature importance on the held-out test set. Unpermuted ensemble MAE was calculated. In addition, it permuted one feature at a time across test observations. Each permutation generated new ensemble predictions. Feature importance represented the increase in MAE after permutation. Each feature underwent 20 permutation repeats using random seed 99. It also evaluated audiometric and clinical features with the same ensemble and test set. Positive importance values indicated greater prediction error after permutation. For clinical evaluation, the analysis exported patient-level predictions. The file included the first-audiogram mean threshold, observed and predicted last-audiogram mean thresholds, observed and predicted changes, absolute prediction error at the last audiogram, and ensemble prediction standard deviation.

### 2.15 Reproducibility and Data Sharing

The analytical pipeline used Python 3.12 with NumPy 2.5.3, pandas 3.0.6, PyTorch 2.14.0, scikit-learn 1.9.1, and SciPy 1.18.1. Jupyter Notebook v7 supported local data processing and computational modelling on a Windows 64-bit system with an Intel i5-1235U CPU and 8 GB RAM. All processing ran offline without shared networks or cloud infrastructure. Predefined random seeds supported reproducibility, and timestamped directories stored model outputs, tuning results, feature importance, predictions, bootstrap results, statistical tests, and a final JSON report. A separate sharing dataset removed patient identifiers and added column-specific Laplace noise to age and audiometric variables. Six noise scales were tested, using 1.0–0.2 for age and 1.0–0.1 for audiometric variables. Five perturbation trials evaluated each scale. The procedure selected the smallest scale with a median Kolmogorov–Smirnov test *p*-value greater than 0.05 and calibrated noise at *α* = 0.05.

### 2.16 Data and Code Availability

The shared dataset excludes patient identifiers and applies statistical perturbation using the Laplace mechanism as a differential privacy approach. Analyses based on the shared dataset may yield results that differ from those reported because of this perturbation. The original dataset may be requested from the researchers. The anonymised dataset and Python scripts used for data processing, statistical analysis, and computational modelling are available on Zenodo: https://doi.org/10.5281/zenodo.23969854.

## 3 Results

Table 1 summarises characteristics of 823 participants. Noise exposure differed by sex, with a higher proportion among females than males (67.9% vs. 26.5%; *p*<0.001). Hypertension was also more frequent among males (54.0% vs. 41.3%; *p*=0.004). Among records with available data for each variable, diabetes was recorded in 79.0% and a positive cholesterol entry in 91.6%; these proportions do not represent population prevalence. Hearing thresholds at the first and last audiograms did not differ significantly between sexes.

**Table 1:**
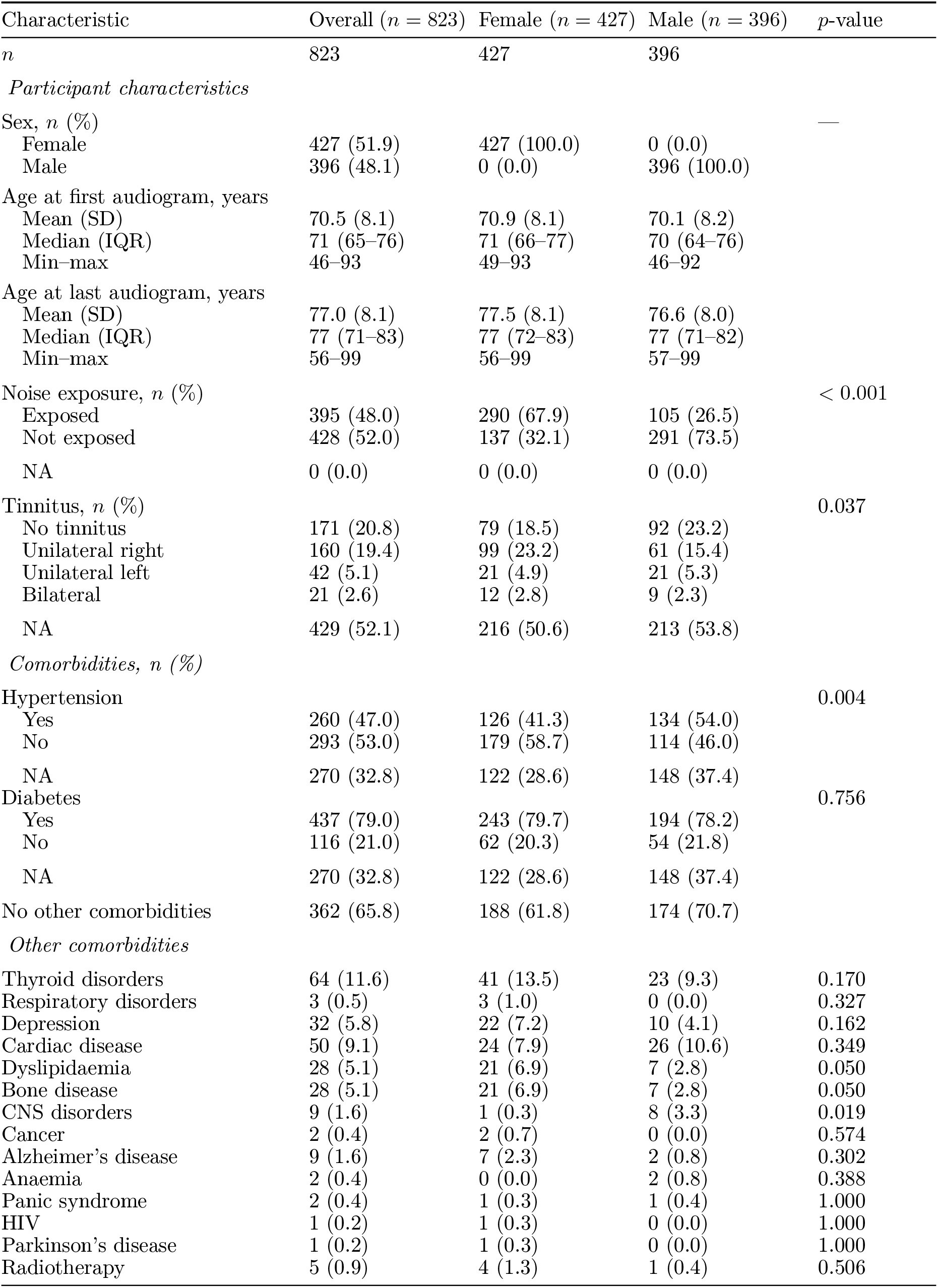
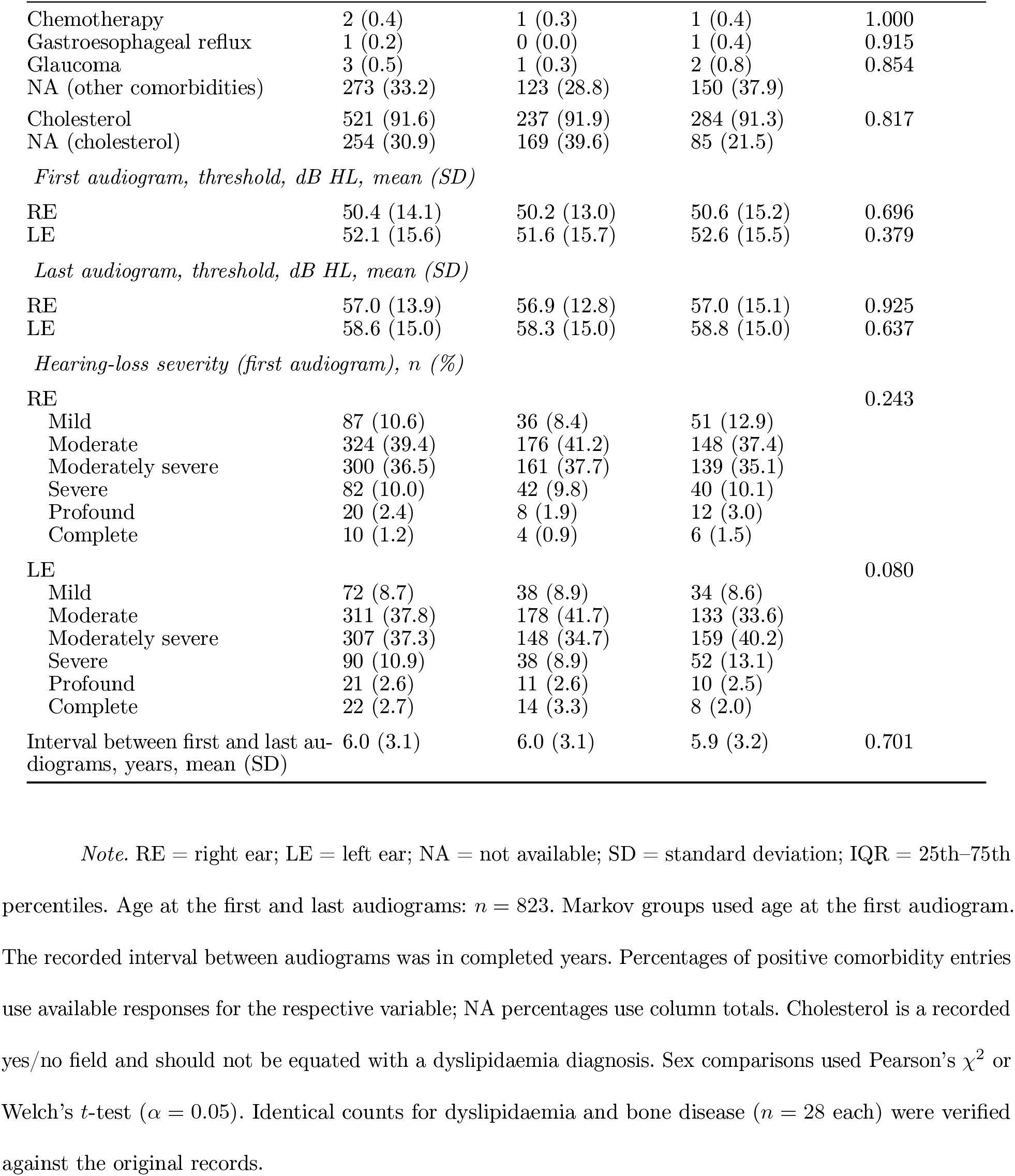
Characteristics of the Study Sample, Overall and by Sex.

| Characteristic | Overall (*n* = 823) | Female (*n* = 427) | Male (*n* = 396) | *p*-value |
|---|---|---|---|---|
| *n* | 823 | 427 | 396 |  |
| *Participant characteristics* |  |  |  |  |
| Sex, *n* (%) |  |  |  | — |
| Female | 427 (51.9) | 427 (100.0) | 0 (0.0) |  |
| Male | 396 (48.1) | 0 (0.0) | 396 (100.0) |  |
| Age at first audiogram, years |  |  |  |  |
| Mean (SD) | 70.5 (8.1) | 70.9 (8.1) | 70.1 (8.2) |  |
| Median (IQR) | 71 (65–76) | 71 (66–77) | 70 (64–76) |  |
| Min–max | 46–93 | 49–93 | 46–92 |  |
| Age at last audiogram, years |  |  |  |  |
| Mean (SD) | 77.0 (8.1) | 77.5 (8.1) | 76.6 (8.0) |  |
| Median (IQR) | 77 (71–83) | 77 (72–83) | 77 (71–82) |  |
| Min–max | 56–99 | 56–99 | 57–99 |  |
| Noise exposure, *n* (%) |  |  |  | < 0.001 |
| Exposed | 395 (48.0) | 290 (67.9) | 105 (26.5) |  |
| Not exposed | 428 (52.0) | 137 (32.1) | 291 (73.5) |  |
| NA | 0 (0.0) | 0 (0.0) | 0 (0.0) |  |
| Tinnitus, *n* (%) |  |  |  | 0.037 |
| No tinnitus | 171 (20.8) | 79 (18.5) | 92 (23.2) |  |
| Unilateral right | 160 (19.4) | 99 (23.2) | 61 (15.4) |  |
| Unilateral left | 42 (5.1) | 21 (4.9) | 21 (5.3) |  |
| Bilateral | 21 (2.6) | 12 (2.8) | 9 (2.3) |  |
| NA | 429 (52.1) | 216 (50.6) | 213 (53.8) |  |
| *Comorbidities, n (%)* |  |  |  |  |
| Hypertension |  |  |  | 0.004 |
| Yes | 260 (47.0) | 126 (41.3) | 134 (54.0) |  |
| No | 293 (53.0) | 179 (58.7) | 114 (46.0) |  |
| NA | 270 (32.8) | 122 (28.6) | 148 (37.4) |  |
| Diabetes |  |  |  | 0.756 |
| Yes | 437 (79.0) | 243 (79.7) | 194 (78.2) |  |
| No | 116 (21.0) | 62 (20.3) | 54 (21.8) |  |
| NA | 270 (32.8) | 122 (28.6) | 148 (37.4) |  |
| No other comorbidities | 362 (65.8) | 188 (61.8) | 174 (70.7) |  |
| *Other comorbidities* |  |  |  |  |
| Thyroid disorders | 64 (11.6) | 41 (13.5) | 23 (9.3) | 0.170 |
| Respiratory disorders | 3 (0.5) | 3 (1.0) | 0 (0.0) | 0.327 |
| Depression | 32 (5.8) | 22 (7.2) | 10 (4.1) | 0.162 |
| Cardiac disease | 50 (9.1) | 24 (7.9) | 26 (10.6) | 0.349 |
| Dyslipidaemia | 28 (5.1) | 21 (6.9) | 7 (2.8) | 0.050 |
| Bone disease | 28 (5.1) | 21 (6.9) | 7 (2.8) | 0.050 |
| CNS disorders | 9 (1.6) | 1 (0.3) | 8 (3.3) | 0.019 |
| Cancer | 2 (0.4) | 2 (0.7) | 0 (0.0) | 0.574 |
| Alzheimer’s disease | 9 (1.6) | 7 (2.3) | 2 (0.8) | 0.302 |
| Anaemia | 2 (0.4) | 0 (0.0) | 2 (0.8) | 0.388 |
| Panic syndrome | 2 (0.4) | 1 (0.3) | 1 (0.4) | 1.000 |
| HIV | 1 (0.2) | 1 (0.3) | 0 (0.0) | 1.000 |
| Parkinson’s disease | 1 (0.2) | 1 (0.3) | 0 (0.0) | 1.000 |
| Radiotherapy | 5 (0.9) | 4 (1.3) | 1 (0.4) | 0.506 |
| Chemotherapy | 2 (0.4) | 1 (0.3) | 1 (0.4) | 1.000 |
| Gastroesophageal reflux | 1 (0.2) | 0 (0.0) | 1 (0.4) | 0.915 |
| Glaucoma | 3 (0.5) | 1 (0.3) | 2 (0.8) | 0.854 |
| NA (other comorbidities) | 273 (33.2) | 123 (28.8) | 150 (37.9) |  |
| Cholesterol | 521 (91.6) | 237 (91.9) | 284 (91.3) | 0.817 |
| NA (cholesterol) | 254 (30.9) | 169 (39.6) | 85 (21.5) |  |
| *First audiogram, threshold, dB HL, mean (SD)* |  |  |  |  |
| RE | 50.4 (14.1) | 50.2 (13.0) | 50.6 (15.2) | 0.696 |
| LE | 52.1 (15.6) | 51.6 (15.7) | 52.6 (15.5) | 0.379 |
| *Last audiogram, threshold, dB HL, mean (SD)* |  |  |  |  |
| RE | 57.0 (13.9) | 56.9 (12.8) | 57.0 (15.1) | 0.925 |
| LE | 58.6 (15.0) | 58.3 (15.0) | 58.8 (15.0) | 0.637 |
| *Hearing-loss severity (first audiogram), n (%)* |  |  |  |  |
| RE |  |  |  | 0.243 |
| Mild | 87 (10.6) | 36 (8.4) | 51 (12.9) |  |
| Moderate | 324 (39.4) | 176 (41.2) | 148 (37.4) |  |
| Moderately severe | 300 (36.5) | 161 (37.7) | 139 (35.1) |  |
| Severe | 82 (10.0) | 42 (9.8) | 40 (10.1) |  |
| Profound | 20 (2.4) | 8 (1.9) | 12 (3.0) |  |
| Complete | 10 (1.2) | 4 (0.9) | 6 (1.5) |  |
| LE |  |  |  | 0.080 |
| Mild | 72 (8.7) | 38 (8.9) | 34 (8.6) |  |
| Moderate | 311 (37.8) | 178 (41.7) | 133 (33.6) |  |
| Moderately severe | 307 (37.3) | 148 (34.7) | 159 (40.2) |  |
| Severe | 90 (10.9) | 38 (8.9) | 52 (13.1) |  |
| Profound | 21 (2.6) | 11 (2.6) | 10 (2.5) |  |
| Complete | 22 (2.7) | 14 (3.3) | 8 (2.0) |  |
| Interval between first and last au-diograms, years, mean (SD) | 6.0 (3.1) | 6.0 (3.1) | 5.9 (3.2) | 0.701 |

*Note.* RE = right ear; LE = left ear; NA = not available; SD = standard deviation; IQR = 25th–75th percentiles. Age at the first and last audiograms: *n* = 823. Markov groups used age at the first audiogram. The recorded interval between audiograms was in completed years. Percentages of positive comorbidity entries use available responses for the respective variable; NA percentages use column totals. Cholesterol is a recorded yes/no field and should not be equated with a dyslipidaemia diagnosis. Sex comparisons used Pearson’s χ<sup>2</sup> or Welch’s *t*-test (*α* = 0.05). Identical counts for dyslipidaemia and bone disease (*n* = 28 each) were verified against the original records.

Table 2 presents the paired comparison between the 1D-CNN and linear regression. The CNN ensemble (test-set evaluation) showed significantly lower absolute error than linear regression in the LE after Holm correction (*p*_adj_ = 0.0328). The RE comparison yielded a non-significant result (*p*_adj_ = 0.1969).

**Table 2:** CNN versus Linear Regression: Paired Comparisons.

| Ear | Mean diff | 95% CI / seed range | *p*-value | *d* |
|---|---|---|---|---|
| *MAE, nested cross-validation* |  |  |  |  |
| RE | 0.422 | [0.242 – 0.574] | — | — |
| LE | -0.123 | [-0.257 – 0.040] | — | — |
| *RMSE, nested cross-validation* |  |  |  |  |
| RE | 0.591 | [0.416 – 0.753] | — | — |
| LE | -0.017 | [-0.256 – 0.173] | — | — |
| *R*<sup>2</sup>*, nested cross-validation* |  |  |  |  |
| RE | -0.047 | [-0.063 – -0.033] | — | — |
| LE | 0.0003 | [-0.012 – 0.014] | — | — |
| *Absolute error, fixed split* |  |  |  |  |
| RE | 0.360 | [-0.191 – 0.886] | 0.197<sup>*</sup> | 0.146 |
| LE | -0.599 | [-1.100 – -0.122] | 0.033<sup>*</sup> | -0.271 |

*Note.* CNN = convolutional neural network; RE = right ear; LE = left ear; MAE = mean absolute error; RMSE = root mean squared error; *R*<sup>2</sup> = coefficient of determination; CI = confidence interval; *d* = standardised effect size. Mean differences represent CNN minus linear regression. Nested cross-validation ranges represent minimum–maximum differences across ten seeds. MAE, RMSE and *R*<sup>2</sup> comparisons were descriptive and were not tested for statistical significance. Fixed-split 95% CIs were obtained using participant-level bootstrap (2,000 resamples). Paired permutation tests used 9,999 permutations. <sup>\*</sup> indicates Holm-adjusted *p*-values. Confidence intervals were not adjusted for multiplicity. Dash (—) indicates no calculated values.

During nested cross-validation, the CNN achieved MAEs of 6.475 dB HL (RE) and 6.047 dB HL (LE), compared with 6.054 dB HL (RE) and 6.170 dB HL (LE) for linear regression (Table 3). The CNN (RE) achieved an *R*^2^ of 0.631, while linear regression achieved 0.678. Similarly, the CNN (LE) and linear regression both achieved an *R*^2^ of 0.72 (rounded to two decimal places). Among audiometric features from the first audiogram, 500 and 4000 Hz showed the largest positive permutation importance in the RE. In the LE, 250, 500 and 2000 Hz showed the largest values. Clinical variables showed smaller effects.

**Table 3:** Predictive Performance and Permutation Feature Importance.

| *Panel A. Predictive performance* |  |  |  |
| Ear | MAE | RMSE | *R*<sup>2</sup> |
|---|---|---|---|
| *Linear regression, nested cross-validation* |  |  |  |
| RE | 6.054 [5.974 – 6.188] | 8.165 [8.048 – 8.292] | 0.678 [0.659 – 0.693] |
| LE | 6.170 [6.039 – 6.312] | 8.376 [8.159 – 8.571] | 0.720 [0.692 – 0.738] |
| *1D-CNN, nested cross-validation* |  |  |  |
| RE | 6.475 [6.359 – 6.613] | 8.756 [8.621 – 8.834] | 0.631 [0.612 – 0.652] |
| LE | 6.047 [5.958 – 6.126] | 8.360 [8.212 – 8.474] | 0.720 [0.697 – 0.738] |
| *Linear regression, fixed split* |  |  |  |
| RE | 6.098 | 8.653 | 0.598 |
| LE | 6.558 | 8.862 | 0.721 |
| *CNN ensemble, fixed split* |  |  |  |
| RE | 6.459 | 8.977 | 0.567 |
| LE | 5.959 | 8.404 | 0.749 |
| *Persistence, fixed split* |  |  |  |
| RE | 8.769 | — | — |
| LE | 8.114 | — | — |
| *Panel B. Permutation feature importance* |  |  |  |
| Feature | RE | LE |  |
| *First audiogram hearing thresholds* |  |  |  |
| 250 Hz | 0.044 | 0.047 |  |
| 500 Hz | 0.095 | 0.028 |  |
| 1000 Hz | 0.024 | 0.012 |  |
| 2000 Hz | 0.042 | 0.028 |  |
| 3000 Hz | 0.050 | 0.009 |  |
| 4000 Hz | 0.053 | 0.019 |  |
| 6000 Hz | 0.049 | 0.005 |  |
| 8000 Hz | 0.016 | -0.007 |  |
| *Clinical variables* |  |  |  |
| Age (first audiogram) | -0.008 | -0.016 |  |
| Sex | -0.002 | -0.022 |  |
| Occupational noise exposure | 0.018 | 0.005 |  |
| Tinnitus | -0.003 | -0.001 |  |
| Hypertension | -0.009 | 0.001 |  |
| Diabetes | -0.026 | -0.018 |  |

*Note.* RE = right ear; LE = left ear; CNN = convolutional neural network; MAE = mean absolute error; RMSE = root mean squared error; *R*<sup>2</sup> = coefficient of determination; CI = confidence interval. Values in square brackets represent descriptive 2.5th–97.5th percentiles across ten seeds, not 95% CIs. MAE and RMSE are expressed in dB HL. Fixed-split results are point estimates from 79 records per ear; CIs were not calculated for these individual metrics. Bootstrap 95% CIs were calculated for paired absolute-error differences (Table 2). The CNN ensemble averaged predictions from eight models. Persistence was evaluated using MAE only. Permutation importance represents change in held-out MAE after permuting one feature. Negative values indicate reduced prediction error. Dash (—) indicates no calculated values.

Markov analysis included 823 observed transitions per ear. In the right ear (RE), 347 (42.2%) showed worsening, 435 (52.9%) remained stable, and 41 (5.0%) showed improvement. Corresponding values for the left ear (LE) were 337 (40.9%), 459 (55.8%), and 27 (3.3%), respectively.

Overall worsening probabilities reached 34.0% for the RE and 35.3% for the LE (Table 4). Hypertension and diabetes showed no significant between-group differences after adjustment. Occupational noise exposure produced a 4.8 percentage-point difference in the RE and a 1.5 percentage-point difference in the LE; neither comparison was statistically significant after Holm adjustment. Worsening also increased across age groups, reaching 43.3% in the RE and 42.0% in the LE among participants aged *≥* 80 years.

**Table 4:**
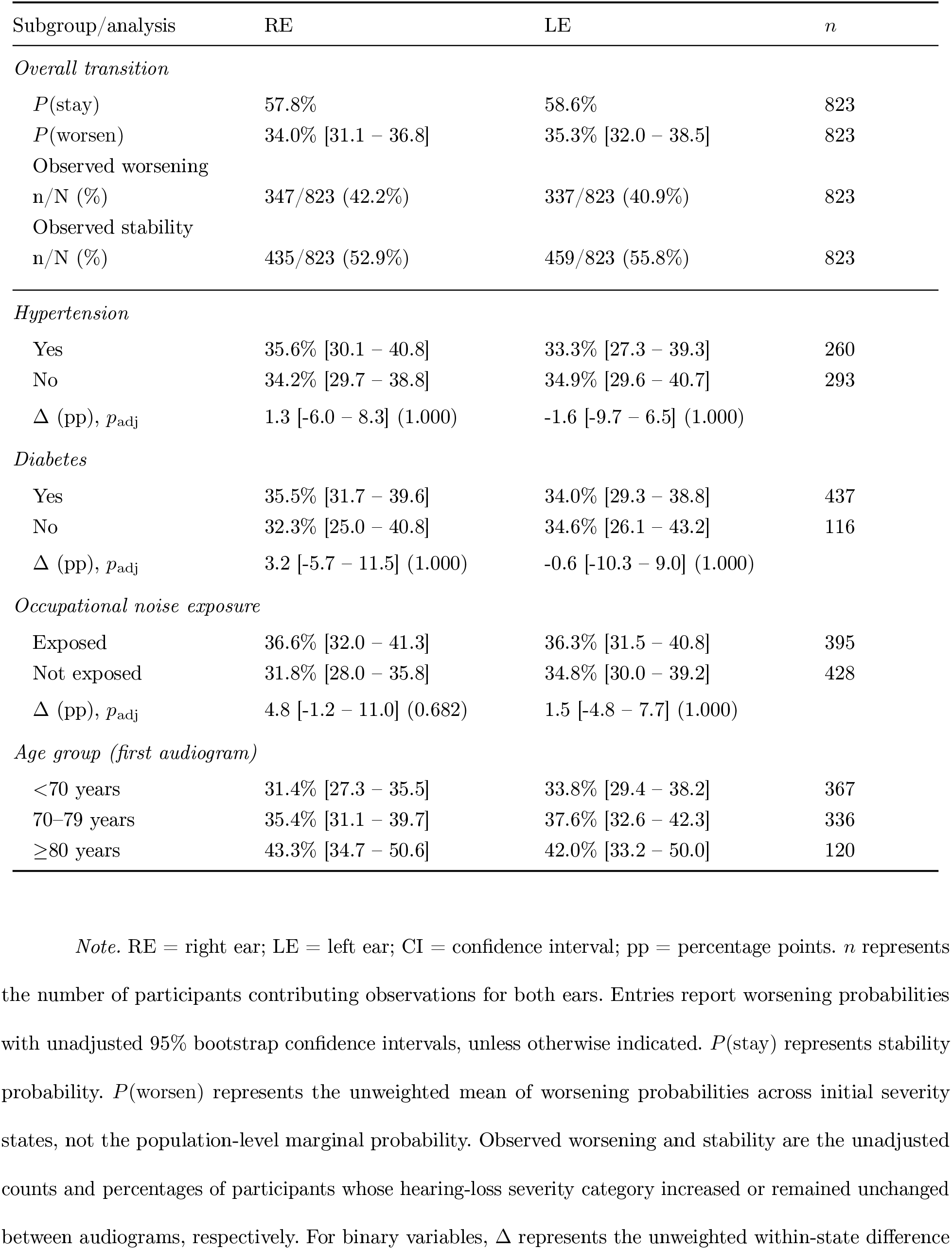
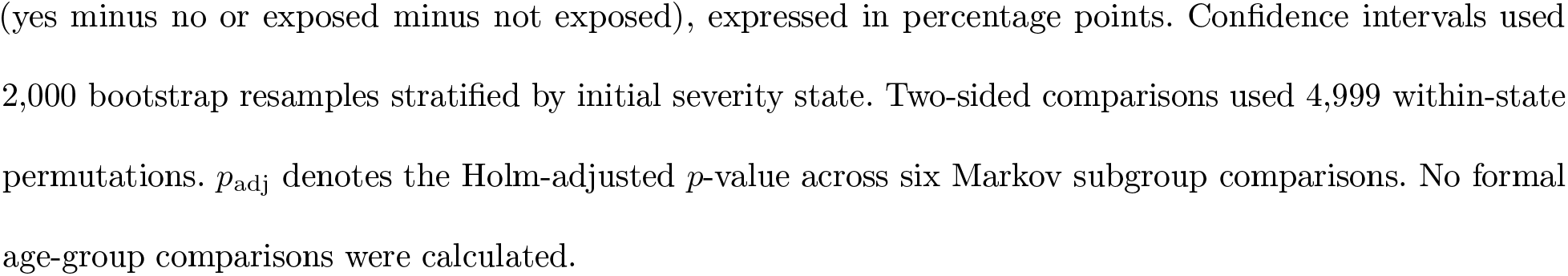
Markov Chain Estimates of Hearing-Loss Worsening.

| Subgroup/analysis | RE | LE | *n* |
|---|---|---|---|
| *Overall transition* |  |  |  |
| *P*(stay) | 57.8% | 58.6% | 823 |
| *P*(worsen) | 34.0% [31.1 – 36.8] | 35.3% [32.0 – 38.5] | 823 |
| Observed worsening<br>n/N (%) | 347/823 (42.2%) | 337/823 (40.9%) | 823 |
| Observed stability<br>n/N (%) | 435/823 (52.9%) | 459/823 (55.8%) | 823 |
| *Hypertension* |  |  |  |
| Yes | 35.6% [30.1 – 40.8] | 33.3% [27.3 – 39.3] | 260 |
| No | 34.2% [29.7 – 38.8] | 34.9% [29.6 – 40.7] | 293 |
| Δ (pp), *p*<sub>adj</sub> | 1.3 [-6.0 – 8.3] (1.000) | -1.6 [-9.7 – 6.5] (1.000) |  |
| *Diabetes* |  |  |  |
| Yes | 35.5% [31.7 – 39.6] | 34.0% [29.3 – 38.8] | 437 |
| No | 32.3% [25.0 – 40.8] | 34.6% [26.1 – 43.2] | 116 |
| Δ (pp), *p*<sub>adj</sub> | 3.2 [-5.7 – 11.5] (1.000) | -0.6 [-10.3 – 9.0] (1.000) |  |
| *Occupational noise exposure* |  |  |  |
| Exposed | 36.6% [32.0 – 41.3] | 36.3% [31.5 – 40.8] | 395 |
| Not exposed | 31.8% [28.0 – 35.8] | 34.8% [30.0 – 39.2] | 428 |
| Δ (pp), *p*<sub>adj</sub> | 4.8 [-1.2 – 11.0] (0.682) | 1.5 [-4.8 – 7.7] (1.000) |  |
| *Age group (first audiogram)* |  |  |  |
| <70 years | 31.4% [27.3 – 35.5] | 33.8% [29.4 – 38.2] | 367 |
| 70–79 years | 35.4% [31.1 – 39.7] | 37.6% [32.6 – 42.3] | 336 |
| ≥80 years | 43.3% [34.7 – 50.6] | 42.0% [33.2 – 50.0] | 120 |

*Note.* RE = right ear; LE = left ear; CI = confidence interval; pp = percentage points. *n* represents the number of participants contributing observations for both ears. Entries report worsening probabilities with unadjusted 95% bootstrap confidence intervals, unless otherwise indicated. *P*(stay) represents stability probability. *P*(worsen) represents the unweighted mean of worsening probabilities across initial severity states, not the population-level marginal probability. Observed worsening and stability are the unadjusted counts and percentages of participants whose hearing-loss severity category increased or remained unchanged between audiograms, respectively. For binary variables, Δ represents the unweighted within-state difference (yes minus no or exposed minus not exposed), expressed in percentage points. Confidence intervals used 2,000 bootstrap resamples stratified by initial severity state. Two-sided comparisons used 4,999 within-state permutations. *p*<sub>adj</sub> denotes the Holm-adjusted *p*-value across six Markov subgroup comparisons. No formal age-group comparisons were calculated.

## 4 Discussion

This study compared a convolutional neural network (CNN) with linear regression to predict hearing thresholds at the last audiogram. Markov chains were used to estimate transitions between hearing-loss severity categories. The cohort comprised older adults with established bilateral hearing loss and recorded clinical information.

Nested cross-validation (performance testing on separate folds) indicated lower CNN prediction error in the left ear (LE). The right ear (RE) showed no improvement. The fixed-test analysis (separate test set evaluation) also favoured the CNN in the LE after Holm correction. Markov estimates differed numerically across ears, age groups, and occupational noise exposure; the occupational noise contrast was not statistically significant. Hypertension and diabetes also showed no statistically significant differences in worsening probabilities.

Earlier applications estimated current hearing thresholds or classified audiograms (Gathman et al., 2023; Kassjański et al., 2024; Song et al., 2015). The present study predicted subsequent hearing thresholds and estimated probabilities of worsening and stability across hearing-loss severity categories.

### 4.1 Predictive performance: CNN compared to linear regression

Linear regression served as a parsimonious benchmark for assessing whether model complexity improved prediction. Both models used identical predictors, data partitions, and seeds, allowing paired comparisons under identical conditions. During nested cross-validation, the CNN had a higher MAE of 6.475 versus 6.054 dB HL in the RE. In the LE, MAE decreased from 6.170 to 6.047 dB HL. The CNN also had a lower *R*^2^ of 0.631 versus 0.678 in the RE, with both models yielding an *R*^2^ of approximately 0.72 in the LE. The lower CNN error in the LE could be considered exploratory, as gains were not consistent across ears and evaluation strategies.

Audiogram structure may have supported the convolutional architecture. Hearing thresholds are arranged from lower to higher frequencies, allowing the CNN to examine values at neighbouring frequencies. These values may capture slopes and variations in audiometric configuration. Convolutional filters can learn local combinations through shared weights, whereas ordinary linear regression assigns additive coefficients to individual inputs. Parallel filters spanning three and five frequencies were used to process patterns at different scales.

Pooling layers then summarised extracted audiometric features before their combination with clinical inputs. Compared with linear regression, the CNN architecture directly incorporates relationships between neighbouring frequencies. The CNN may therefore represent non-linear relationships without specifying interactions in advance. Related work used CNN-based feature extraction to estimate hearing thresholds from otoacoustic emissions (Liu & Gong, 2024). However, otoacoustic emission measurements differ from pure-tone audiograms.

### 4.2 Model Choice

The present study did not compare other machine learning algorithms for predicting hearing thresholds. Random forests, support vector regression, or other neural architectures could identify different patterns in the same data. The findings therefore cannot establish superiority of this CNN over other machine learning methods for predicting hearing thresholds. In a related investigation, Chen et al. (2024) applied machine learning to predict hearing thresholds over time in individuals with genetic sensorineural hearing loss. The long short-term memory model achieved an MAE of 4.34 dB HL. This value was lower than CNN MAEs observed in the present study (5.959–6.475 dB HL). Differences in population characteristics, observation periods, and prediction methods limit direct performance comparisons. Both investigations nevertheless suggest that machine learning may contribute to longitudinal prediction of hearing thresholds using previous audiometric information.

Model choice may also depend on the structure of available observations. Recurrent networks are designed to process sequences, whereas the present dataset included two assessments per participant. Longer audiometric series might provide additional information for sequence-based models.

### 4.3 Differences between ensembles in the fixed split

The CNN ensemble showed lower prediction errors than linear regression in the LE when evaluated on a separate test set (fixed-test analysis). The CNN did not show lower errors in the RE. The ensemble achieved an MAE of 6.459 dB HL (RE), compared with 6.098 dB HL (RE) for linear regression. Corresponding LE values were 5.959 and 6.558 dB HL.

Paired permutation comparisons identified a significant difference in LE absolute error after Holm correction (*p*_adj_ = 0.0328). The difference in the RE was not statistically significant (*p*_adj_ = 0.1969). Thus, the LE advantage was also present in the separate test set.

Differences between ears may partly reflect differences in evaluation procedures. Nested cross-validation assessed performance across five outer folds and ten seeds. A separate sample was used to evaluate averaged predictions of eight CNNs (held-out test). Variations in sample composition and averaging of model predictions may have contributed to the observed discrepancy. In this sense, data do not establish a consistent predictive advantage of the CNN over linear regression in both ears across evaluation strategies.

### 4.4 Relevance of audiometric and clinical variables

The largest increase in MAE occurred at 500 Hz (RE; 0.095 dB HL) and 250 Hz (LE; 0.047 dB HL). Small increases suggest limited sensitivity of model predictions to individual frequencies. Permutation importance reflects changes in prediction error rather than hearing deterioration over time.

Linssen et al. (2014) reported an association between poorer initial hearing thresholds and greater deterioration over 12 years. The longitudinal association reported by Linssen et al. (2014) differs from frequency-specific predictive importance measured in the present study. The present analysis therefore cannot establish whether frequencies with greater predictive importance also undergo greater deterioration.

Regarding clinical predictors, permutation analysis in the present study produced smaller changes in MAE, including small negative permutation-importance estimates for diabetes in both ears. This pattern may indicate limited contributions of these variables to ensemble predictions. Low predictive importance does not exclude possible associations with hearing progression.

The model processed audiometric and clinical information through separate structures. The pipeline arranged eight hearing thresholds from the first audiogram in fixed frequency order. A second branch processed age, sex, tinnitus, hypertension, diabetes, and noise exposure. The fusion layer then combined these representations to predict subsequent thresholds. This structure preserved audiometric ordering while incorporating clinical information. In addition, standardisation supported training with variables measured on different scales. The CNN processed adjacent audiometric frequencies as neighbouring inputs, although intervals between frequencies differed in hertz.

### 4.5 Progression of hearing loss using Markov chains

Markov chains estimated mean conditional probabilities of worsening of 34.0% (RE) and 35.3% (LE). Corresponding probabilities of stability were 57.8% (RE) and 58.6% (LE). These estimates are unweighted means across initial severity states and differ from observed proportions. Observed worsening affected 347/823 (42.2%) (RE) and 337/823 (40.9%) (LE); observed stability affected 435/823 (52.9%) (RE) and 459/823 (55.8%) (LE).

Similarly, Vaden et al. (2017) examined changes in audiometric classifications over time. Audiometric phenotypes remained stable in 61.5% of ears despite age-related deterioration in hearing thresholds. For comparison with that observed proportion, the proportions remaining in the same severity category were 52.9% (RE) and 55.8% (LE) in the present study. These studies used different classifications: audiometric phenotypes in Vaden et al. (2017) and hearing-loss severity in the present study. The outcomes may therefore not be directly comparable, despite both being observed proportions of stability.

Changes in hearing thresholds and transitions between severity categories may describe different aspects of hearing loss progression. Thresholds may worsen without crossing a severity boundary. Small changes near a boundary may alter classification. Thus, transitions between categories do not directly measure the magnitude of threshold deterioration. This distinction also limits comparisons across studies using different severity classifications or intervals between audiograms.

Regarding age, estimated worsening probabilities increased across age groups. Among participants aged 80 years or older, worsening probabilities reached 43.3% in the RE and 42.0% in the LE. Linssen et al. (2014) also reported an association between increasing age and greater hearing deterioration between audiograms. These studies assessed different outcomes, and age-group differences were not formally tested in the present analysis.

For occupational noise exposure, the Markov analysis estimated a difference of 1.5 percentage points in worsening probability in the LE. The RE showed a difference of 4.8 percentage points. Neither difference was statistically significant after Holm adjustment. Gopinath et al. (2021) associated workplace noise exposure with hearing loss incidence over ten years but found no association with progression. Dillard, Humes, et al. (2025) reported associations between noise exposure history and rates of hearing threshold change across frequencies. The same study found no differences in overall pure-tone average progression between exposure groups.

The Markov analysis found no statistically significant differences in worsening probabilities for hypertension or diabetes. Mick et al. (2023) reported an association between hypertension and hearing deterioration over approximately three years. Cross-sectional analyses in that study also identified an association between diabetes and hearing thresholds. Differences in observation intervals, outcome definitions, and analytical methods may partly explain contrasting findings.

Although none of the subgroup contrasts reached statistical significance, the five-state Markov analysis quantified conditional worsening and stability across severity categories. Estimates provide a descriptive account of categorical change that complements continuous threshold prediction.

### 4.6 Implications

CNN estimated subsequent hearing thresholds in dB HL using first-audiogram and clinical information. Markov chains estimated probabilities of stability and worsening across hearing-loss severity categories. These approaches characterised longitudinal hearing changes through continuous predictions and categorical transitions.

Changes in hearing thresholds do not necessarily result in transitions between severity categories. Similarly, transitions between categories do not quantify the magnitude of hearing threshold changes. CNN predictions and Markov probabilities may therefore provide complementary information about longitudinal hearing changes.

The interpretation of these estimates also differs. Test-set MAE describes prediction error of the CNN. Markov transition probabilities summarise changes across initial severity states rather than individual prognoses.

### 4.7 Limitations

Retrospective design and records from a single hearing health service may limit generalisability. Audiometric examinations were documented at the same hearing health service, and hearing-loss severity was classified using World Health Organization criteria. Assessments covered 2008 to 2023, with a mean recorded interval of 6.0 years between audiograms. Clinical records contained missing information, and clinical covariates were not imputed. Incomplete information on occupational noise intensity and duration may affect subgroup comparisons. Excluding records without tinnitus information may have reduced the predictive sample and could introduced selection bias. The resulting models were not evaluated in participants without documented tinnitus information.

Audiometric imputation was fitted within training partitions to reduce information leakage. Repeated cross-validation across seeds reused participants from the same dataset and could not replace external validation. Ear-specific audiometric profiles, outcome variability, or sampling variability in the held-out cohort may explain the CNN’s advantage in the LE but not the RE. These explanations remain hypotheses because differences in model effects between ears were not formally tested.

Neither the CNN nor the Markov analysis explicitly modelled time between audiograms; predictions refer to observed last-audiogram thresholds, and Markov estimates are not annual transition rates. Transition probabilities were averaged with equal weight across initial severity states, regardless of participant counts. These estimates therefore describe conditional transitions within the study population rather than individual risks over time.

Further studies could evaluate model performance and reproducibility in other populations. Comparisons between convolutional and alternative models could use identical training-fitted preprocessing and examine performance across severity at the first audiogram. Larger cohorts could also support time-specific Markov transitions, incorporate time between audiograms, and assess calibration within initial severity states. Such comparisons may establish whether greater model complexity provides reproducible benefits.

### 4.8 Conclusions

CNN showed lower prediction error than linear regression in the LE during nested cross-validation. Evaluation on a separate test set also favoured the CNN in the LE. No statistically significant difference was observed in the RE. Markov chains estimated probabilities of worsening and stability across hearing-loss severity categories. Worsening probabilities increased numerically across age groups. No statistically significant differences were observed for occupational noise exposure, hypertension, or diabetes. This characterisation may support assessment of changes in hearing thresholds alongside the likelihood of progression to more severe hearing-loss categories. The results describe an exploratory ear-specific CNN advantage and quantify transitions between hearing-loss severity categories. Reported findings may contribute to understanding longitudinal hearing changes through distinct outcomes.

## Acknowledgements

The authors thank the patients whose clinical records contributed to this study. We also acknowledge the Hearing Health Service of the Speech-Language Pathology and Audiology Clinic, University of São Paulo, Bauru, Brazil, for its support and contribution to this research.

## Declarations

### Ethics approval and consent to participate

The Research Ethics Committee of the Bauru School of Dentistry, University of São Paulo, approved the study under protocol no. 1,745,659, in accordance with Resolution 466/2012 of the Brazilian National Health Council.

### Data availability

A privacy-protected version of the study dataset is available on Zenodo: https://doi.org/10.5281/zenodo.23969854. The shared dataset excludes patient identifiers and includes statistical perturbation using the Laplace mechanism as a differential privacy approach. Variations in reported results may occur during independent executions due to randomness introduced by statistical perturbation. The original dataset may be requested from the researchers.

### Use of AI-assisted tools

Proton Lumo was used to assist with language revision and refactoring of author-developed analysis code. The authors reviewed all AI-assisted changes and retain full responsibility for the analyses, code, and manuscript content and integrity.

### Code availability

Python scripts used for data processing, statistical analysis, and computational modelling are available on Zenodo: https://doi.org/10.5281/zenodo.23969854.

### Author contributions

Conceptualization: HGCM and KAPC contributed to conception and design of the study. Data curation: HGCM and LJ were responsible for data curation. Investigation: HGCM and SRTG conducted the investigation. Methodology: HGCM, KAPC, and LJ developed the study methodology. Formal analysis: HGCM and KAPC performed formal analysis of data. Software: HGCM developed the software used in the study. Validation: KAPC performed validation of results. Supervision: KFA and LJ supervised development of the study. Writing – original draft: HGCM, SRTG, and LJ prepared the initial version of the manuscript. Writing – review and editing: All authors contributed to reviewing and editing the manuscript. This manuscript has been seen and approved by all listed authors.

